# Elevated TRAF6 expression confers radioresistance and predicts poor prognosis in cervical cancer

**DOI:** 10.64898/2026.07.09.26357625

**Authors:** Jiongyu chen, Yingming Jin, Hongcheng Li, Xinqi Lv, Qihao Zhao, Zebiao Ma, Yongkang Yang, Dong-Hua Yang, Li Zhou, Lin Peng

## Abstract

**Background:** The lack of effective biomarkers and therapeutic targets to overcome radioresistance in cervical cancer remains a major clinical challenge. Tumor necrosis factor receptor-associated factor 6 (TRAF6), an E3 ubiquitin ligase pivotal in immune and inflammatory signaling, has been implicated in various malignancies. However, its role in radioresistance in cervical cancer remains unclear.

**Methods:** TRAF6 expression was evaluated in cervical cancer tissues from 162 patients who underwent postoperative radiotherapy at our institution and in 304 cases from the TCGA-CESC cohort. The prognostic significance of TRAF6 was assessed using Kaplan-Meier and Cox regression analyses. A nomogram integrating TRAF6 expression with clinicopathological factors was constructed to predict overall survival (OS) and progression-free survival (PFS). The functional role of TRAF6 in malignant phenotypes and radiosensitivity was investigated using shRNA-mediated knockdown in HeLa and C33A cervical cancer cells. Immune cell infiltration patterns associated with TRAF6 expression were analyzed using ssGSEA and xCELL algorithms based on TCGA data.

**Results:** TRAF6 expression was significantly elevated in cervical cancer tissues compared with adjacent normal tissues (70.99% vs. control, *P* < 0.001) and was higher in radioresistant than in radiosensitive patients (*P* < 0.001). High TRAF6 expression was associated with shorter OS (HR = 18.73, *P* = 0.004) and PFS (HR = 8.44, *P* < 0.001) and was identified as an independent risk factor for radiotherapy resistance (OR = 8.44, *P* < 0.001). The TRAF6-integrated nomogram demonstrated good predictive accuracy for OS (C-index = 0.7351) and PFS (C-index = 0.7444). TRAF6 knockdown in cervical cancer cells significantly suppressed proliferation, migration, and invasion, while substantially enhancing radiosensitivity of tumor cells. Functional enrichment analysis revealed that TRAF6-related genes were enriched in autophagy, mitophagy, and HPV infection pathways. Immune cell infiltration analysis showed that TRAF6 expression correlated with distinct immune cell profiles, characterized by enrichment of activated dendritic cells, M1 macrophages, and regulatory T cells, alongside depletion of cytotoxic effectors such as CD8+ T cells and γδ T cells.

**Conclusions:** TRAF6 could be a prognostic biomarker associated with poor outcomes and indicator of radiotherapy resistance in cervical cancer, TRAF6 represents a potential therapeutic target for overcoming radioresistance in cervical cancer.

## 1 Introduction

Cervical cancer presents a substantial global health challenge for women, ranking as the fourth most common cause of cancer incidence and mortality among women globally. Although organized cervical screening programs and universal prophylactic human papillomavirus (HPV) vaccination have markedly reduced cervical cancer incidence, absolute global mortality burden remains substantial, especially in low- and middle-income countries^[1]^. Radiotherapy is the cornerstone of treatment for advanced cervical cancer, and irradiation technologies have advanced dramatically over the past few decades^[2]^. However, clinical outcomes are still suboptimal due to uncontrolled primary tumor progression, locoregional recurrence and distant metastasis driven by intrinsic or acquired resistance to ionizing radiation^[3]^. Therefore, the identification of predictive biomarkers for radiotheapeutic response is urgently needed to enable risk-stratified patient management and to serve as potential therapeutic targets for radiosensitization strategies.

Radioresistance in cervical cancer arises through a multifactorial interplay of molecular mechanisms, including hypoxia-inducible factor 1-alpha (HIF-1α) signaling activation, reactive-oxygen species production, enhanced DNA damage repair, tumor microenvironment remodeling, metabolic reprogramming, expansion of cancer stem cells, and deregulation of cell cycling or apoptosis^[4]^. Increasing evidence implicates that gene alterations in cervical carcinogenesis can substantially reshape cellular behavior and the surrounding tumor microenvironment, thereby critically driving cancer progression and metastasis. The genomic perturbations are frequently associated with distinct molecular expression patterns, among which modifications in protein level represent the most functionally informative biological indicators for therapeutic stratification^[5]^.

Tumor necrosis factor receptor-associated factor 6 (TRAF6), a RING domain-containing E3 ubiquitin ligase that orchestrates immune responses and inflammatory signaling, is highly expressed across multiple human malignancies and recognized as an oncogene^[6]^. TRAF6 overexpression has been correlated with advanced tumor stage and unfavorable survival outcomes in breast cancer^[7]^, colon cancer^[8]^, and gastric cancer^[9]^, underscoring its potential utility as both a predictive biomarker and a therapeutic target^[10]^. Mechanistically, TRAF6 exerts a significant role in enhancing malignant phenotypes of tumor cells by mediating hypoxia signaling and inflammatory cascades, stimulating autophagy, regulating metabolic reprogramming, and remodeling the immunosuppressive microenvironment^[10, 11]^. In cervical cancer cells, TRAF6 was found to confer tumor cell growth, metastasis^[12]^, and paclitaxel resistance^[13]^. Intriguingly, a recent study indicated that gold nanoparticles enhance radiosensitivity in glioma cells through suppression of TRAF6/NF-κB-mediated CCL2 secretion, underscoring TRAF6 as a promising molecular target for overcoming radioresistance^[14]^. Nevertheless, the predictive value of TRAF6 for radioresistance and its underlying mechanisms remains unknown.

In this study, we first determined TRAF6 expression and evaluated its prognostic significance in cervical cancer specimens from patients who received radiotherapy, using a cohort from our hospital and data from publicly available transcriptomic datasets. Subsequently, we established a TRAF6-based nomogram integrating TRAF6 expression with clinicopathological variables for predicting prognosis before radiotherapy. In addition, we validated the functional role of TRAF6 in malignant progression and radioresistance of cervical cancer cells by siRNA-mediated TRAF6 silencing. Finally, we investigated the potential mechanisms of TRAF6 by analyzing relevant gene pathway enrichment and immune cell infiltration using TCGA datasets. Our findings identify TRAF6 as a novel prognostic biomarker and a potential therapeutic target for overcoming radioresistance in cervical cancer, offering a new strategy to enhance radiosensitivity and improve long-term patient survival.

## 2 Materials and Methods

### 2.1 Tissue Specimen collection, clinical data acquisition, and patient follow-up

Tumor specimens and corresponding clinical data were collected from cervical cancer patients who underwent surgical resection followed by adjuvant radiotherapy at the Cancer Hospital of Shantou University Medical College between January 2015 and December 2022. All patients provided written informed consents for the research use of their specimens and clinical data. Cases were included based on the following criteria: (i) pathologically confirmed cervical cancer and postoperative radiotherapy performed at our institution; and (ii) complete clinical, imaging, and follow-up data available. Exclusion criteria comprised: (i) preoperative radiotherapy or chemotherapy; (ii) prior cervical surgery or hysterectomy; (iii) incomplete cervix; (iv) concurrent or history of other malignancies; (v) pregnancy at diagnosis; and (vi) active inflammatory disease of the reproductive tract. Clinicopathological parameters, including age, menopause status, body mass index (BMI), disease history, pathological type, FIGO stage, primary tumor size, and invasion/infiltration degree, were obtained from medical records. Additionally, samples of histologically confirmed normal adjacent cervical tissue were collected as the controls. The follow-up endpoint was set at January 2025. This study was approved by the Ethics Review Committee of the Affiliated Cancer Hospital of Shantou University Medical College (Approval No: 2026-KY-08).

### 2.2 Immunohistochemistry (IHC) and evaluation

IHC staining for TRAF6 was performed on formalin-fixed, paraffin-embedded tissue sections using a standardized protocol^[15]^. Primary antibody targeting TRAF6 (1:200 dilution, #592Y, Abcam, USA) were applied, along with corresponding secondary antibody (Fuzhou Maixin Biotechnology Ltd). The results of IHC staining were independently evaluated by two board-certified pathologists who were blinded to clinical outcomes, employing a semi-quantitative scoring system. Staining intensity was graded as: 0 (negative), 1 (weak), 2 (moderate), and 3 (strong). The proportion of positive tumor cells was assessed using a four-tier system: 1 (<25%), 2 (25–50%), 3 (51–75%), and 4 (76–100%). A final staining index (range: 0–12) was calculated by multiplying the intensity and proportion scores. The optimal cutoff value for stratifying patients into TRAF6-low (scores 0–3) and TRAF6-high (scores 4–12) expression groups was determined using X-tile software (version 3.6.1) based on survival outcomes.

### 2.3 Public data acquisition and functional gene enrichment analysis

RNA-sequencing data and matched clinical annotations for cervical squamous cell carcinoma and endocervical adenocarcinoma (CESC) were retrieved from The Cancer Genome Atlas (TCGA) database (https://portal.gdc.cancer.gov/). The Gene Expression Profiling Interactive Analysis (GEPIA2) was used to obtain the expression of key genes in tumor and normal samples from various human tissues^[16]^. Samples were divided into low- and high-TRAF6 expression groups based on the median transcripts per million (TPM). Differentially expressed gene (DEG) expression analysis was carried out using the limma package in R (v4.4.2), with gene annotation supported by org.Hs.eg.db, clusterProfiler, enrichplot, ggplot2, and ggrepel. Functional enrichment of TRAF6-associated genes was investigated through Gene Ontology (GO) biological process and Kyoto Encyclopedia of Genes and Genomes (KEGG) pathways. Enrichment significance was determined by |NES| > 1, false discovery rate (FDR) < 0.25, and adjusted *P*-value < 0.05.

### 2.4 Immune cell infiltration analysis

The relationship between TRAF6 expression and the tumor immune microenvironment in cervical cancer was analyzed using RNA-seq data from the TCGA-CESC cohort. The relative abundances of infiltrating immune cells were estimated using multiple computational algorithms, including CIBERSORT (v1.03), xCell, and ssGSEA. TRAF6 expression was dichotomized into high and low groups using the maxstat R package (v0.7-25) based on the optimal cutoff value identified through maximally selected rank statistics with respect to survival. Differences in immune cell infiltration between TRAF6-high and TRAF6-low groups were assessed by Wilcoxon rank-sum tests, with Benjamini-Hochberg correction for multiple comparisons (FDR < 0.05). The results of immune cell correlation were visualized using ggplot2 (v3.3.5) and pheatmaps (v1.0.12). The analytical scripts are publicly accessible at GitHub (github.com/username/repo), ensuring full reproducibility of the results using TCGA-CESC dataset.

### 2.5 Nomogram construction and validation

We conducted univariate and multivariate Cox proportional hazards regression analyses to assess the prognostic significance of TRAF6 expression and clinicopathological variables for overall survival (OS) and progression-free survival (PFS). Subsequently, a nomogram model was constructed as practical predictive tools for individualized 1-, 3-, and 5-year OS and PFS. The predictive accuracy of the nomograms was evaluated using calibration curves. Model discrimination was quantified by the consistency index (C-index).

### 2.6 Cell culture and treatment

Human cervical cancer cell lines HeLa (CRM-CCL-2, ATCC) and C33A (TCHu176, Shanghai Cell Bank, Chinese Academy of Sciences) were maintained in RPMI 1640 medium and RPMI DMEM medium (SH30809.01B, SV30303.01, HyClone), respectively. Complete growth media were supplemented with 10% fetal bovine serum (FBS, Gibco, USA), 1% penicillin-streptomycin (Gibco, USA). Cells were incubated at 37 °C in a humidified atmosphere with 5% CO₂. For irradiation, cells were exposed to 6 MV X-rays delivered by a linear accelerator (23EX; Varian, United States) at single doses of 0, 2, 4, 6, 8, and 10 Gy. Irradiation parameters were set as follows: field size, 30 × 30 cm; source-to-surface distance, 100 cm; and dose rate, 285 cGy/min.

### 2.7 Gene silencing using shRNA lentiviral transduction

To knock down TRAF6 expression, cells were transduced with recombinant lentiviral particles encoding TRAF6-targeting short hairpin RNAs (shRNAs). The following encoded shRNA sequences were used: shTRAF6#1: 5΄-TTAGAGAGGTCACTTACTATT-3΄ and shTRAF6#2: 5΄-GCCACGGGAAATATGTAATAT-3΄. A non-targeting control shRNA (shNC: 5΄-TTCTCCGAACGTGTCACGTTT-3΄) was used in parallel. Briefly, cells were seeded at a density of 5 × 10⁴ cells per well in 6-well plates and allowed to adhere for 24 hours. Subsequently, lentiviral transduction was performed at multiplicity of infection (MOI) 20 in medium supplemented with 5 μg/mL polybrene. Stable transductants were selected via puromycin treatment (500 ng/mL) initiated 48 hours post-transduction.

### 2.8 Cell viability assay

Cell viability was assessed using the Cell Counting Kit-8 (CCK-8, Beyotimy, China) according to the manufacturer’s instructions. Cervical cancer (CC) cells were seeded in 96-well plates at a density of 1 × 10⁴ cells per well and cultured overnight at 37°C in a 5% CO₂ incubator. At designated time points (0, 24, 48 or 72 hours post-irradiation), 10 µL of CCK-8 reagent was added to each well, followed by incubation for 3 hours at 37°C. Absorbance at 450 nm was measured using a microplate reader (MK3, Thermo, USA). All experiments were performed in biological triplicate with technical duplicates.

### 2.9 Wound healing assay

Cell migratory capacity was evaluated using a wound healing assay. Cells were cultured in 12-well plates until reaching full confluence. A standardized linear scratch was then made using a 20 µL pipette tip, and the cells were incubated in RPMI 1640 medium for up to 72 hours. Images of the same wound areas were captured at 0, 24, 48 and 72 hours post-scratching. Wound closure was quantified as percentage of initial wound area using ImageJ software.

### 2.10 Cell invasion assay

Cell invasion was assessed using Transwell chambers (BIOFIL, Guangzhou, China) with 8 µm pore size membranes pre-coated with Matrigel. The lower compartment was filled with 500 µL of complete medium (RPMI 1640 medium containing 10% FBS) as chemoattractant. A total of 1 × 10⁵ cells in 200 µL serum-free RPMI 1640 medium supplemented with 1% BSA were seeded into the upper chamber. Following 48-hour incubation, the upper chambers were removed. Invaded cells on the upper surface were fixed, stained with 0.1% crystal violet, and enumerated in five randomly selected fields per membrane under a light microscope (DM3000, Leica, Germany) at 40× magnification.

### 2.11 Clonogenic cell survival assay

Following irradiation at specific doses, cells were cultured for 14 days to allow colony formation. Colonies were fixed in 4% paraformaldehyde, stained with Giemsa (Sigma, USA) for 10 minutes, and enumerated microscopically. Colonies containing ≥50 cells were quantified to calculate surviving fractions. Surviving fraction (SF) was determined by colonies formed / cells seeded. The sensitization enhancement ratio (SER) was determined from cell survival curves based on the multi-target/single-hit model.

### 2.12 Western blotting

Protein concentration was quantified using a BCA Protein Assay Kit (G3522, GBCBIO). Equal amounts of protein (50 µg) were resolved by 10% SDS-polyacrylamide gel electrophoresis and subsequently transferred onto a polyvinylidene fluoride (PVDF) membrane (IPVH00010, Millipore). Primary antibodies used were as follows: rabbit anti-vinculin (1:1000, S0B1132, Starter-BIO) and rabbit anti-TRAF6 (1:1000, #592Y, Abcam). Corresponding HRP-conjugated secondary antibodies were goat anti-mouse (1:5000, #7076S, CST) and goat anti-rabbit (1:5000, #7074S, CST).

### 2.13 Statistical analysis

We conducted statistical analyses using SPSS (version 24.0) and GraphPad Prism (version 9.0), and visualized in R (version 4.4.2). For continuous variables, optimal cutoff values were determined using X-tile software (3.6.1, Yale) based on survival outcomes. Categorical variables were compared using chi-square, and continuous variables using Student’s t-test or Mann-Whitney U test as appropriate. Based on the Response Evaluation Criteria for Solid Tumors (RECIST), patients experiencing recurrence or progression within 12 months after radiotherapy were classified as the radiotherapy-resistant group, while those without recurrence within one year were classified as the radiotherapy-sensitive group. Logistic regression analysis was performed to identify risk factors for radioresistance. Variables achieving *P* < 0.10 in univariate analysis were eligible for entry into multivariate analysis using forward stepwise selection. Results are presented as odds ratios (ORs) with 95% confidence intervals (CIs). OS was measured from hospital admission to death from any cause, while PFS was defined as the time from radiotherapy initiation to the first documented recurrence or death from any cause. Patients alive without events were censored at their last follow-up before this cutoff date. Survival distributions were estimated using the Kaplan-Meier analysis and compared by log-rank tests. Univariate and multivariate Cox proportional hazards regression models were subsequently conducted to identify independent prognostic factors. All statistical tests were two-sided, and *P* < 0.05 were considered statistically significant.

## 3 Results

### 3.1 Expression levels of TRAF6 in CESC tumor tissues and adjacent tissues

A total of 162 cervical cancer samples and 39 matched adjacent normal cervical tissues from our institutional cohort were analyzed. Immunohistochemistry staining revealed that TRAF6 was predominantly localized in the cytoplasm and plasma membrane of tumor cells. Notably, TRAF6 was either absent or only faintly detectable in normal cervical epithelium, whereas a marked elevation was observed in tumor tissues (70.99%, 115/162; *P* < 0.001, Figure 1A-B). To explore TRAF6 expression at the transcriptional level, we interrogated the TCGA and GTEx databases using the GEPIA2 platform. Unexpectedly, no statistically significant difference in TRAF6 mRNA levels was found between tumor and normal tissues (*P* > 0.05, Figure 1C). These findings suggest that the elevated TRAF6 protein expression in cervical cancer may be regulated at the post-transcriptional level.

**Figure 1.**
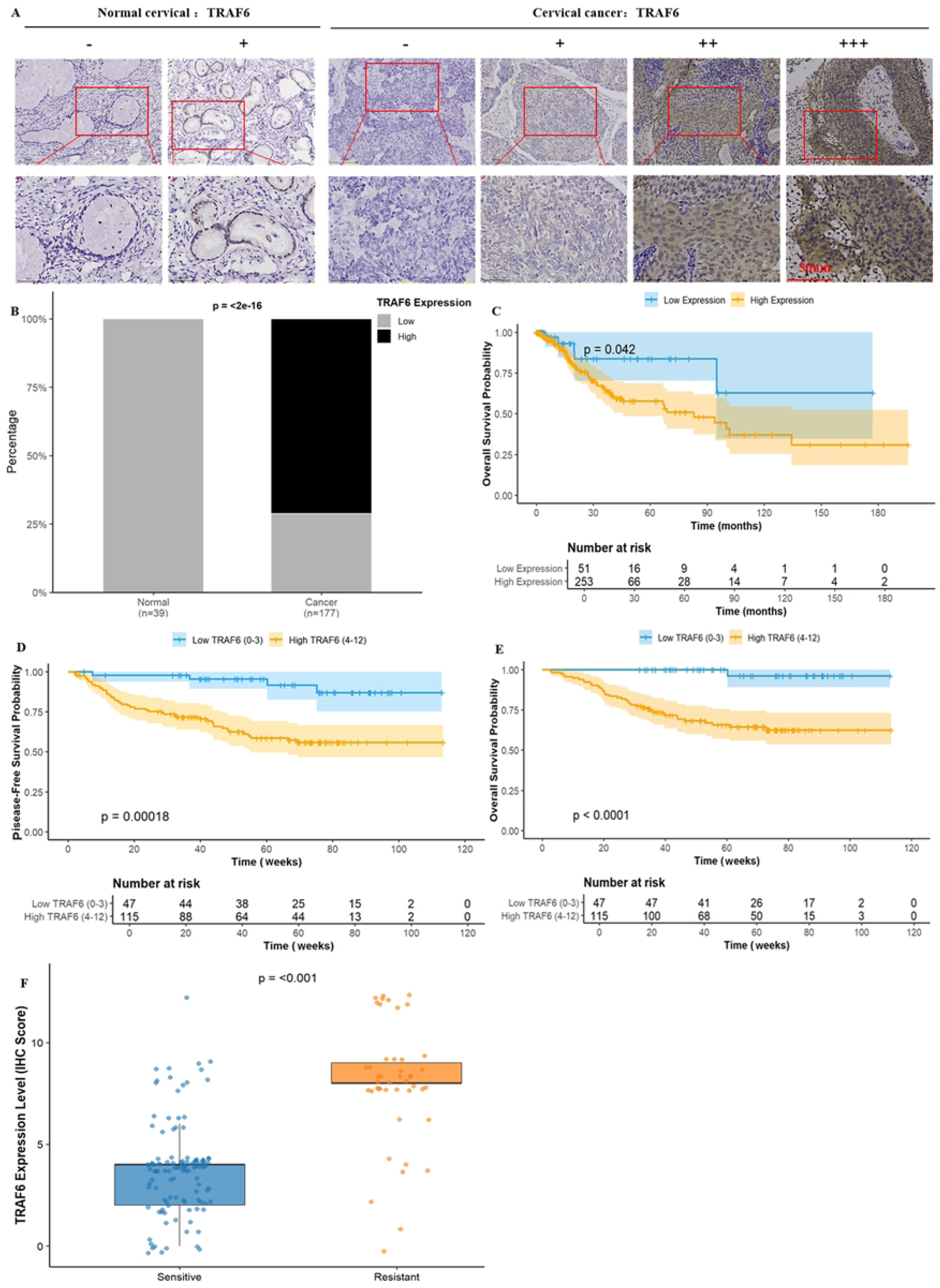
Upregulation of TRAF6 expression correlates with poor prognosis of cervical cancer. (A) Representative TRAF6-negative and +, ++, +++ immunostaining in normal cervical tissues and cervical cancer tissues. (B) Expression and distribution of TRAF6 in normal cervical tissues and cervical cancer tissues. (C) Kaplan-Meier analysis of OS based on TRAF6 expression levels in the TCGA-CESC cohort. Kaplan-Meier curves for OS(D) and DFS(E) in the institutional cohort stratified by TRAF6 expression. (F) Comparison of TRAF6 expression levels between radiosensitive and radioresistant group.

### 3.2 Elevated expression of TRAF6 in tumor specimens correlates with radioresistance and poor prognosis

The data of a total of 304 patients with cervical cancer from the TCGA cohort were collected and categorized into high- and low-TRAF6 expression groups. Kaplan–Meier survival analysis demonstrated that high TRAF6 expression was significantly associated with poor OS (HR = 1.73, *P* = 0.042, Figure 1D). Furthermore, TRAF6 expression levels were significantly higher in the radioresistant group compared to the radiosensitive group (*P* < 0.001, Figure 1G). Survival analysis indicated that elevated TRAF6 expression was associated with shorter OS (HR = 18.73, *P* = 0.004, Figure 1E) and PFS (HR = 8.44, *P* < 0.001, Figure 1F). These findings collectively underscore that elevated TRAF6 expression is associated with adverse prognosis in cervical cancer patients undergoing postoperative radiotherapy.

### 3.3 Prognostic factors for survival and radiotherapy Outcomes

Cox proportional-hazards regression was performed to identify prognostic determinants of PFS and OS in our cohort. Univariate analysis demonstrated that age >58 years (HR = 2.16), hypertension (HR = 3.53), high TRAF6 expression (HR = 18.73), and distant metastasis (HR = 12.90) were associated with worse OS. Additionally, age >58 years (HR = 8.41), hypertension (HR = 2.53), high TRAF6 expression (HR = 5.66), and parametrial invasion (HR = 2.86) were associated with shorter PFS (all *P* < 0.05). Multivariate analysis further identified age >58 years (HR = 18.73), hypertension (HR = 16.55), and distant metastasis (HR = 16.14) as independent prognostic factors for adverse OS (all *P* < 0.001) (Table 2), whereas age >58 years (HR = 5.66), hypertension (HR = 5.15) and parametrial invasion (HR = 5.53) were confirmed as independent predictors of adverse PFS (all *P* < 0.005) (Table 3). To identify risk factors for poor radiotherapy response, univariate logistic regression analysis indicated that hypertension (OR = 2.82), high TRAF6 expression (OR = 8.44), cervical stromal invasion (OR = 7.87), and parametrial invasion (OR = 3.38) were associated with adverse clinical outcomes (all *P* < 0.05). Multivariate logistic regression analysis retained all these factors as independent risk factors (hypertension: OR = 2.82; high TRAF6 expression: OR = 8.44; cervical stromal invasion: OR = 7.87; parametrial invasion: OR = 3.38). (Table 4)

### 3.4 Construction and validation of a nomogram for survival prediction

To enable individualized survival prediction for cervical cancer patients receiving postoperative radiotherapy, we developed a nomogram incorporating the independent prognostic factors identified by multivariable Cox regression. As a result, age, hypertension, TRAF6 expression, TRAF6 expression level and parametrial invasion contributed substantially to the nomogram model, demonstrating strong predictive ability for both OS (C-index = 0.7351) (Figure 2A) and PFS (C-index = 0.7444) (Figure 2C). Calibration plots for 1-, 3-, and 5-year survival rates showed excellent agreement between nomogram-predicted and observed outcomes for both OS (Figure 2B) and PFS (Figure 2D).

**Figure 2.**
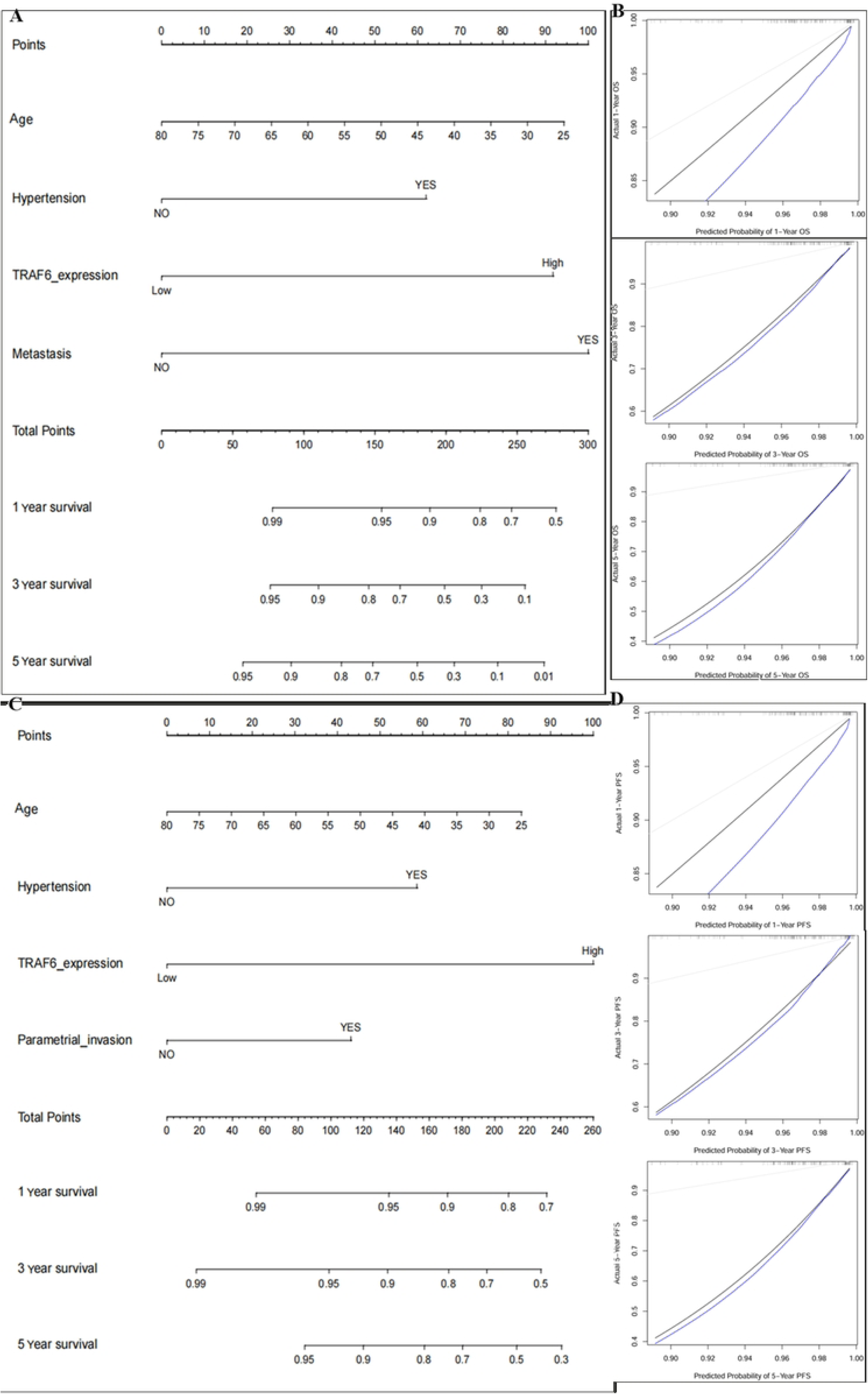
Nomogram and calibration curves for predicting survival and radiotherapy response in cervical cancer patients. (A) Nomogram incorporating TRAF6 expression for predicting OS. (B) Calibration curves for 1-, 3-, and 5- year OS predicted by the nomogram. (C) Nomogram incorporating TRAF6 expression for predicting PFS. (D) Calibration curves for 1-, 3-, and 5- year PFS predicted by the nomogram.

In addition, we investigated the association between TRAF6 expression and radiotherapy response (Table 5). As a result, elevated TRAF6 expression was significantly correlated with radiotherapy resistance (*P* < 0.001). Clinicopathologically, comorbid hypertension (*P* = 0.008), cervical stromal invasion (*P* = 0.021), and parametrial invasion (*P* = 0.024) were independent predictors of reduced radiotherapy sensitivity. No significant associations were observed between radiotherapy response and other variables, including age, BMI, diabetes, histologic type, FIGO stage, tumor size, menopausal status, pelvic lymph node metastasis, or vaginal/bladder invasion status.

### 3.5 Down-regulation of TRAF6 inhibits the proliferation, migration and invasion abilities of cervical cancer HeLa and C33A cells

To ensure stable TRAF6 knockdown, cervical cancer cells were transduced separately with lentiviruses encoding two shRNAs (shTRAF6#1 and shTRAF6#2) that target TRAF6 mRNA. Western blotting confirmed the successful knockdown of TRAF6 (Figure 3A). CCK8 experiments showed that TRAF6 knockdown reduced cervical cancer cell viability (Figure 3B). The scratch wound healing test confirmed that the migration of cervical cancer cells, following TRAF6 knockdown, decreased (Figure 3C), and cell invasion assays showed that cell invasion also declined (Figure 3D).

**Figure 3.**
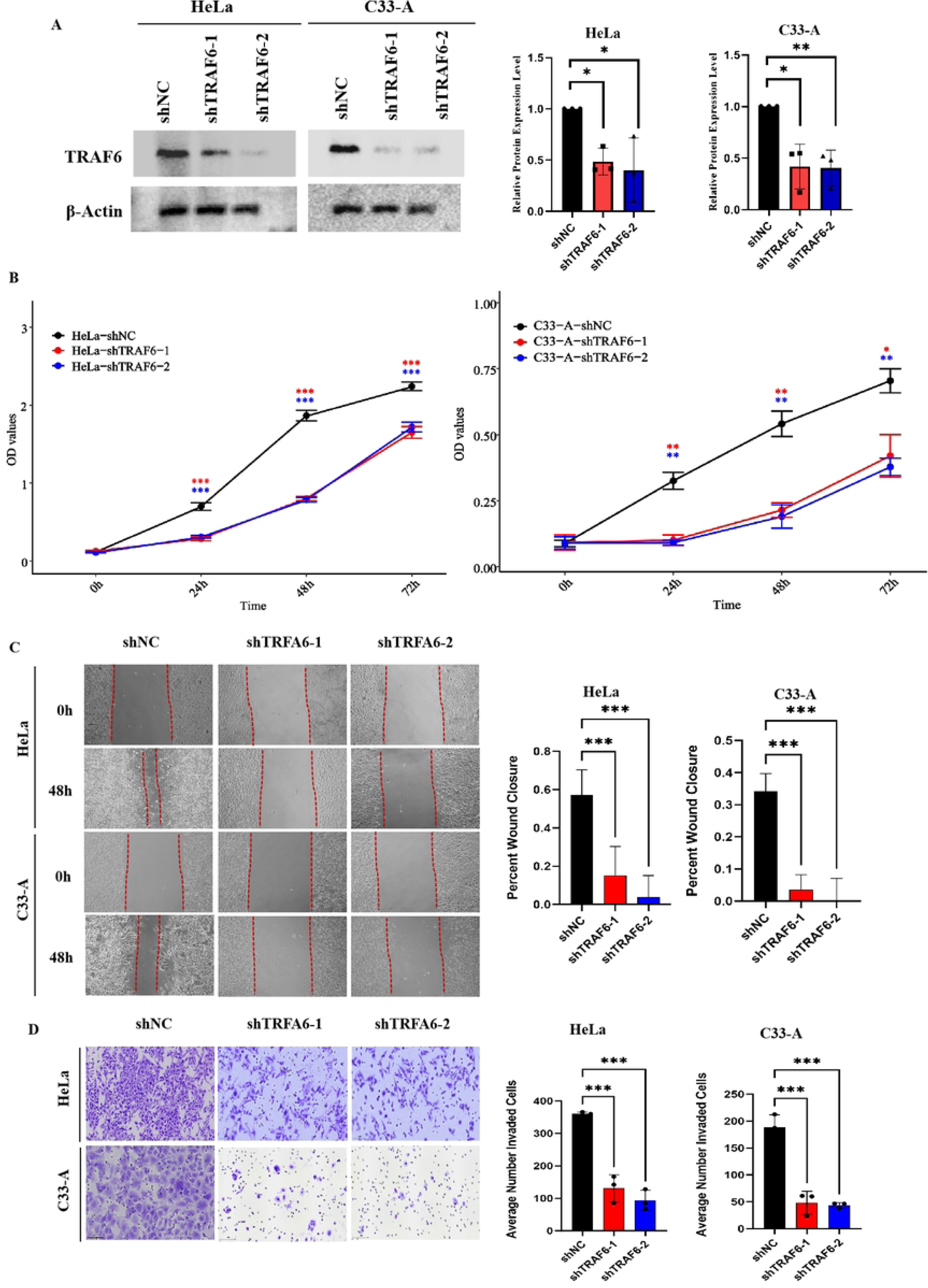
The impact of TRAF6 on proliferation, migration, and invasion of cervical cancer cells. (A) Western blot analysis confirming TRAF6 knockdown efficiency in HeLa and C33A cells using two specific shRNAs (shTRAF6#1 and shTRAF6#2) . (B) CCK-8 assays showing that TRAF6 knockdown significantly suppressed cell proliferation. Wound healing and Transwell invasion assays demonstrating that TRAF6 depletion inhibits cell migration (scale bar = 100 μm; C) and invasion (scale bar = 100 μm; D). Data are presented as mean ± SD from three independent experiments. \**P* < 0.05; \*\**P* < 0.01; \*\*\**P* < 0.001

### 3.6 TRAF6 knockdown enhances radiosensitivity of cervical cancer cells

To validate the role of TRAF6 in radiotherapy of cervical cancer, we evaluated the clonogenic survival of TRAF6-depleted cervical cancer cells following exposure to ionizing radiation. Colony formation assays showed that shTRAF6-transfected HeLa and C33A cells exhibited significantly reduced surviving fractions compared to controls across a range of irradiation doses (0, 2, 4, 6, 8, and 10 Gy) (*P* < 0.01, Figure 4A). Survival curves were fitted using the multi-target single-hit model, from which radiobiological parameters were derived. TRAF6 knockdown resulted in a marked decrease in the quasi-threshold dose (Dq) and mean lethal dose (D0), indicative of enhanced radiosensitivity (*P* < 0.01, Figure 4B-C). These findings suggest that knockdown of TRAF6 sensitizes cervical cancer cells to radiotherapy.

**Figure 4.**
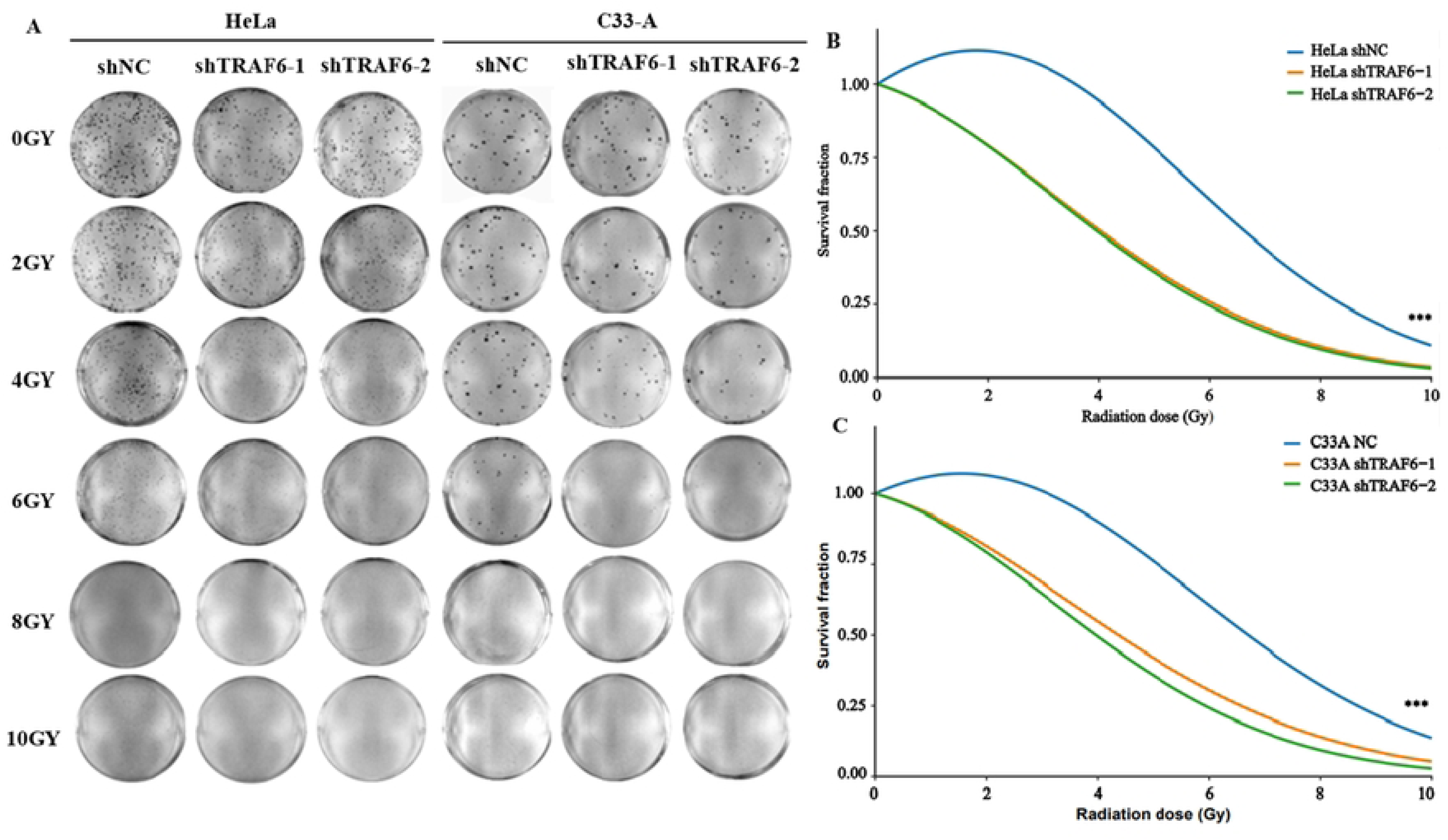
TRAF6 knockdown enhances radiosensitivity of cervical cancer cells. (A) Representative images of colony formation assays in HeLa and C33A cells with or without TRAF6 knockdown following irradiation at indicated doses (0, 2, 4, 6, 8, and 10 Gy). Survival curves fitted using the multi-target single-hit model showing the radiosensitizing effect of TRAF6 depletion in HeLa (B) and C33A cells (C). Data are presented as mean ± SD from three independent experiments. \**P* < 0.05; \*\**P* < 0.01; \*\*\**P* < 0.001.

### 3.7 Functional enrichment analysis of TRAF6-related genes in cervical cancer

To elucidate the potential biological functions of TRAF6 in cervical cancer, we performed pathway enrichment analysis based on TRAF6-related DEGs from the TCGA-CESC datasets. A total of 7193 DEGs were identified between high- and low-TRAF6 expression groups, comprising 548 downregulated and 6,645 upregulated genes. KEGG pathway enrichment analysis showed that these genes were significantly associated with multiple pathways, including human papillomavirus infection, endocytosis, autophagy, mitophagy, cell cycle, protein processing in endoplasmic reticulum and others (Figure 5A). GO analysis delineated their functional roles across three categories. In the Biological Process (BP) domain, genes were primarily enriched in RNA splicing, chromosome segregation, and nucleocytoplasmic transport. For Cellular Component (CC), significant terms included chromosomal region, spindle, and focal adhesion. Regarding Molecular Function (MF), the most notable enrichments were for ubiquitin-like protein transferase activity, protein serine/threonine kinase activity, and GTPase binding (Figure 5B).

**Figure 5.**
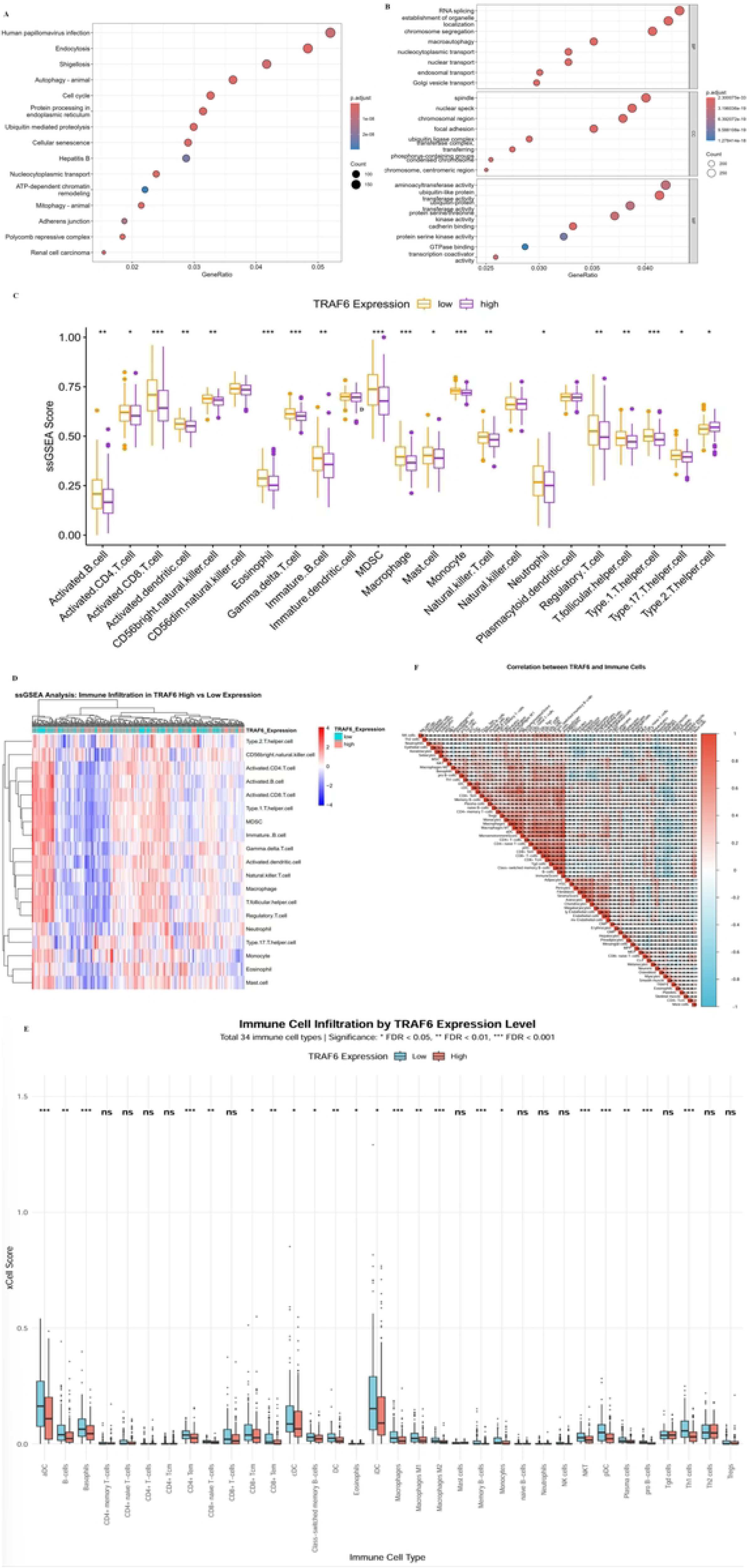
Functional enrichment of TRAF6-associated genes and correlation with immune infiltration. KEGG (A) and GO (B) analyses for the top 7193 TRAF6-related genes in cervical cancer. (C) Overview of TRAF6 expression and immune cell infiltration patterns for the cervical cancer cohort in TCGA. (D) Heatmap of immune cell infiltration analyzed by ssGSEA algorithm. (E) Boxplot of immune cell infiltration levels assessed by the xCELL algorithm. (F) Correlation analysis between TRAF6 expression and immune cell infiltration.

### 3.8 Association between TRAF6 expression and immune cell infiltration

To elucidate the relationship between TRAF6 expression and the tumor immune microenvironment (TIME) in cervical cancer, we employed multiple algorithms to comprehensively analyze the TCGA cohort. As illustrated in Figure 5 C and D, ssGSEA revealed distinct immune cell infiltration patterns. Specifically, the high-TRAF6 expression group exhibited significantly reduced enrichment scores for several critical antitumor immune cells, including activated dendritic cells (aDCs), activated CD8+ T cells, gamma delta (γδ) T cells, and CD56 bright natural killer (NK) cells (*P* < 0.05). Additionally, markers of immune cell activation such as type 17 T helper (Th17) cells and activated CD4+ T cells, were also negatively correlated with TRAF6 levels (*P* < 0.05). Notably, while the overall adaptive immune response appeared suppressed, a shift to the T helper cells was observed, with Th2 cells being relatively enriched compared to Th1 cells (*P* < 0.05). Conversely, innate immune cell populations such as macrophages, myeloid-derived suppressor cells (MDSCs), and immature B cells, also showed a negative association with TRAF6 expression (*P* < 0.05). To further validate these findings, we employed the xCell algorithm to characterize the tumor immune microenvironment associated with TRAF6 expression (Figure 5E and F). Consistently, the high-TRAF6 group exhibited significantly decreased infiltration of aDCs, macrophages, CD8+ central memory T cells (Tcm), and CD8+ effector memory T cells (Tem) and CD4+ Tem (*P* < 0.05). Moreover, the infiltration levels of M1 macrophages, which are classically activated and possess antitumor activity, were significantly lower in TRAF6-high tumors (*P* < 0.01). Similarly, the levels of Th1 cells, which are crucial for cell-mediated antitumor immunity, were significantly reduced in tumors with high TRAF6 expression (*P* < 0.01).

## 4 Discussion

In this study, we investigated the role of TRAF6 in cervical cancer progression and radioresistance through integrating clinical specimen analysis, public database mining, and in vitro experiments. Our institutional cohort demonstrates that TRAF6 protein is significantly upregulated in tumor tissues compared with adjacent normal tissues. Moreover, elevated TRAF6 expression was significantly correlated with poor prognosis in cervical cancer patients receiving postoperative radiotherapy. In vitro study, TRAF6 knockdown in cervical cancer cells significantly attenuated cell proliferation, migration, and invasion, and concurrently enhanced sensitivity to radiotherapy. These findings provide the first evidence, to our knowledge, linking TRAF6 expression to radiosensitivity in solid tumors. Collectively, our results highlight TRAF6 as both a prognostic indicator for patient stratification and a potential therapeutic target for radiosensitization in cervical cancer.

Notably, the upregulation of TRAF6 protein in tumor specimens was not recapitulated at the mRNA level, indicating a potential post-transcriptional regulatory mechanism. These observations align with previous reports that TRAF6 expression can be modulated via protein stabilization^[17]^, deubiquitination^[18]^, or microRNA-mediated translational control^[19]^. The discordance between protein and mRNA levels warrants further investigation into the role of post-translational modifications and degradation pathways in TRAF6 regulation during cervical carcinogenesis. The oncogenic role of TRAF6 has been well established in multiple solid tumors, including gastric cancer ^[20]^, colorectal cancer ^[21]^ and lung cancer ^[22]^. In cervical cancer, prior studies have implicated TRAF6 in tumor growth, metastasis, and paclitaxel-resistance^[12, 13]^. In line with these reports, TRAF6 silencing in HeLa and C33A cells led to attenuated malignant phenotypes and enhanced radiosensitivity. However, no significant correlation was found between TRAF6 levels and conventional clinicopathological parameters such as FIGO stage, tumor size, or depth of invasion. This apparent discrepancy may be attributed to several factors. First, clinicopathological staging systems such as FIGO primarily reflect anatomical tumor extent and macroscopic morphological features rather than the molecular heterogeneity of individual tumors. The prognostic value of a biomarker may thus be independent of, or even orthogonal to, traditional staging parameters. Second, the tumor microenvironment comprises complex, multifactorial interactions among stromal components, immune cell infiltration, and metabolic regulation, which collectively determine clinical outcomes and may obscure the isolated contribution of a single molecule ^[12]^. In this context, TRAF6 may exert its oncogenic effects through context-dependent signaling networks that are not fully captured by conventional histopathological assessment. In addition, the relatively homogeneous distribution of TRAF6 overexpression across different stages may limit its discriminatory power for stratifying anatomical progression, while still retaining prognostic significance for survival outcomes.

Further analyses based on the TCGA-CESC dataset and our institutional cohort consistently demonstrated that higher TRAF6 mRNA and protein levels were significantly associated with shorter OS. In addition, TRAF6 protein expression was also positively correlated with radioresistance and reduced PFS. The TRAF6-integrated nomogram model for OS and PFS prediction yielded C-indices of 0.7351 and 0.7444 respectively, indicating good predictive accuracy. This nomogram provides a convenient and effective tool for individualized survival risk assessment in clinical practice. Overall, TRAF6 functions as a key driver of malignant progression and radiotherapy resistance in cervical cancer, underscoring its potential utility as both a prognostic biomarker and a candidate therapeutic target.

The functional gene enrichment analysis provides critical insights into the potential mechanisms through which TRAF6 regulates cervical cancer pathogenesis and radiosensitivity. The enrichment of human papillomavirus (HPV) infection pathway is particularly noteworthy, given that HPV16 oncogenes E6/E7 have been implicated in mediating radiosensitivity by inducing cell cycle arrest^[23]^. As a key adaptor molecule in the TLR/IL-1R signaling cascade, TRAF6 has been shown to modulate inflammatory responses^[24]^, raising the possibility that TRAF6 may influence HPV-mediated radiosensitivity through perturbation of host immune surveillance. Our findings are consistent with previous studies indicating that TRAF6 expression can be upregulated upon HPV stimulation and interacted with HPV16 E6^[25, 26]^. A functional crosstalk between TRAF6 and HPV signaling in cervical cancer radioresistance warrants validation in future studies.

The concurrent enrichment of genes associated with autophagy, mitophagy, and endocytosis pathways suggests that TRAF6 promotes radioresistance in cervical cancer cells by orchestrating cellular homeostasis and stress adaptation. Autophagy has emerged as a dual modulator of radiosensitivity, which confers cytoprotection under moderate stress conditions, whereas excessive activation induces autophagic cell death or sensitizes tumor cells to ionizing radiation^[27]^. Notably, TRAF6-mediated ubiquitination of Beclin-1 has been implicated in autophagy initiation^[28]^, raising the possibility that TRAF6 modulates autophagic flux to influence the intrinsic radiosensitivity of cervical cancer. While mitophagy activation has been implicated as a driver of adaptive radioresistance in cancer cells^[29]^, the involvement of TRAF6 in mitophagy regulation within malignancies remains largely unexplored. Interestingly, TRAF6 has been reported to amplify endoplasmic reticulum stress and disrupt mitochondrial quality control through aberrant mitophagy modulation in hypoxic pulmonary hypertension^[30]^. Moreover, TRAF6 integrates innate immune signals to regulate glucose homeostasis via both Parkin-dependent and Parkin-independent mitophagy pathways^[31]^. Collectively, these findings suggest mitophagy as a potential effector mechanism underlying TRAF6-induced radioresistance in cervical cancer.

Our comprehensive immunogenomic analysis reveals that elevated TRAF6 expression establishes a profoundly immunosuppressive tumor microenvironment in cervical cancer, characterized by the coordinated depletion of both innate and adaptive antitumor immune cell populations. This finding repositions TRAF6 not merely as a signaling adaptor but as a pivotal orchestrator of immune evasion mechanisms, with significant implications for radioresistance and patient stratification.

The marked reduction of aDCs in TRAF6-high tumors represents a critical bottleneck in antitumor immunity. As professional antigen-presenting cells, aDCs bridge innate pathogen sensing and adaptive T cell responses through MHC-peptide complex formation and co-stimulatory molecule upregulation^[32]^. The reduction of this cell subset in the context of TRAF6 overexpression suggests impaired recognition of immunogenic cell death and diminished neoantigen cross-presentation, thereby crippling the priming phase of the cancer-immunity cycle. The suppression of activated CD8⁺ T cells, γδ T cells, and CD56bright NK cells in TRAF6-high tumors reflects a multi-pronged assault on cellular cytotoxicity. For instance, CD8+T cells constitute the principle cytotoxic effectors against cancer in antitumor immunity^[33]^. Their attenuated activation status, coupled with the specific reduction of CD8⁺ Tcm and Tem cell subsets, indicates that TRAF6 disrupts the differentiation and maintenance of tumor-reactive T cell pools. These findings align with prior observations that TRAF6 inhibition enhanced the cytolytic activity of CD8+ T cells^[34]^, further supporting a T cell-intrinsic suppressive function of TRAF6 signaling.

The observation that M1 macrophages are specifically decreased in TRAF6-high tumors, while M2 macrophages and MDSCs show overall negative association with TRAF6, reveals nuanced myeloid compartment remodeling. This pattern diverges from simplistic M1/M2 dichotomy models and instead suggests a broader impairment of myeloid cell fitness and functional polarization induced by TRAF6. Previous research has demonstrated that Th1/Th2 and Th17/Treg imbalances in cervical cancer drive immunological imbalances that intensifies with disease progression^[35]^. The Th1/Th2 paradigm shift observed in our analysis, characterized by Th1 cell reduction and relative Th2 enrichment, suggests TRAF6 may promote a classical immune deviation associated with tumor progression and poor prognosis. Collectively, our findings support a model wherein TRAF6 overexpression in cervical cancer establishes an “immune desert” phenotype characterized by defective antigen presentation, diminished cytotoxic effector recruitment, impaired memory formation, and deviated helper T cell responses, implying the “immune exclusion” or “immunologically ignorant” tumor immune profile.

Despite the important insights provided by this study, several limitations warrant acknowledgment. First, although both the TCGA cohort and our institutional cohort were incorporated, the protein-level validation was performed exclusively using samples from a single center, which may introduce institution-selection biases. Thus, validation in larger, multi-institutional cohorts is needed to ensure the generalizability of our findings. Second, the precise molecular mechanisms through which TRAF6 confers radioresistance remain to be fully elucidated, particularly in the *in vivo* context. Although we demonstrated a functional role for TRAF6 in regulating radiotherapy sensitivity *in vitro*, further mechanistic investigations are required, such as identifying specific ubiquitination targets of TRAF6 in the DNA damage response and clarifying its precise regulatory role in autophagy and mitophagy. Third, our immune cell infiltration analyses were based on bioinformatics predictions derived from public databases and lacked experimental validation in clinical specimens. Consequently, complementary approaches, such as multiplex immunohistochemistry or flow cytometry, are needed to substantiate these findings at the protein and cellular levels. Fourth, the unexpected association between hypertension and clinical outcomes, while adjusted for in multivariate models, may be confounded by unmeasured variables such as antihypertensive medication use, duration of treatment, or the presence of cardiovascular comorbidities. As such, these findings should be interpreted with caution.

## 5. Conclusion

In conclusion, our study establishes the predictive value of TRAF6 in cervical cancer, particularly among patients receiving postoperative radiotherapy, and highlights its potential as a therapeutic target for overcoming radiotherapy resistance. Given the limitations of single-center protein validation and the need for mechanistic elucidation, future multi-center clinical validation, as well as in vitro and in vivo mechanistic investigations, are warranted to translate these findings into clinical practice.

## Ethics statement

The study was approved by the Institute Research Ethics Committee of the Cancer Hospital of Shantou University Medical College (2026-KY-018).

## Author Contributions

Jiongyu Chen: Cell Function Experiment, Immunohistochemical experiment, data curation, software, writing – original draft. Yingming Jin: Cell Function Experiment, software, writing – review & editing. Hongcheng Li: methodology, resources, software. Xinqi Lv: Immunohistochemical experiment, methodology, resources, software. Qihao Zhao: writing – review & editing. Zebiao Ma: funding acquisition, resources. Yongkang Yang: data curation, methodology. Dong-Hua Yang: writing – review & editing. Li Zhou: conceptualization, project administration, writing – review & editing. Lin Peng: conceptualization, project administration, supervision, validation, visualization, writing – review & editing.

## Data Availability

The transcriptomic and clinical data from The Cancer Genome Atlas (TCGA-CESC) and the Genotype-Tissue Expression (GTEx) project used in this study are publicly available via the GDC Data Portal (https://portal.gdc.cancer.gov/) and the GTEx Portal (https://gtexportal.org/home/). All raw data supporting the findings of this study, including the immunohistochemistry scoring matrix, clinicopathological annotations, raw western blot images, and complete cell viability, migration, and invasion assay datasets, are provided in the Supporting Information files accompanying this manuscript. All R scripts used for data analysis and figure generation have been deposited in GitHub (https://github.com/username/repo) and are publicly accessible. The corresponding author can also provide additional materials upon reasonable request.

## Acknowledgments

We extend our deepest respect to all the volunteers for participating in this study and thank Dr Stanley Lin for his constructive comments and language editing.

